# Diagnostic Accuracy of a Smartphone-Enabled Urinary Albumin-to-Creatinine Ratio Test for Detecting Chronic Kidney Disease Among At-Risk Populations

**DOI:** 10.1101/2025.01.20.25320142

**Authors:** Danielle Jeddah, Nicholas B Bevins, Matan Ronen, Navdeep Tangri

**Affiliations:** Healthy.io; BirdRock Laboratories; University of Manitoba, Max Rady College of Medicine

**Keywords:** Chronic Kidney Disease (CKD), Diabetes, Albuminuria, Albumin-to-Creatinine Ratio (uACR), CKD Screening, CKD prevention, Clinical Diagnostics, Point of Care Testing Systems, Medical Device, Diagnostic accuracy, Laboratory Methods and Tools, Machine Learning, KDIGO guidelines, Early detection

## Abstract

**Objective:** To evaluate the diagnostic accuracy of the FDA-cleared Minuteful Kidney Test (MKT), a home-based semi-quantitative urine albumin-to-creatinine ratio (uACR) test, compared to the Beckman Coulter AU 480 analyzer, a standard laboratory quantitative method, for detecting albuminuria in individuals with or at risk for chronic kidney disease (CKD).

**Methods:** Urine samples were obtained from individuals with risk factors for kidney damage. Each sample was analyzed for uACR using both the MKT and the laboratory standard method. Albuminuria was classified according to KDIGO guidelines: A1 (uACR ≤30 mg/g, “Normal to mildly increased”), A2 (uACR 30-300 mg/g, “Moderately increased”), and A3 (uACR ≥300 mg/g, “Severely increased”). Sensitivity, specificity, PPV, and NPV of MKT were calculated against the laboratory standard.

**Results:** Out of 615 collected urine samples, 555 met inclusion criteria. Albuminuria (uACR ≥30 mg/g) was detected in 24.9% of samples using the laboratory method (95% CI, 21.3–28.5%). The MKT demonstrated a sensitivity of 96.4%, specificity of 84.2%, NPV of 98.6%, and PPV of 66.8%. The test accurately categorized 85.6% of samples into the KDIGO albuminuria categories A1, A2, and A3, accurately identifying 100% of laboratory-confirmed A3 samples as abnormal.

**Conclusions:** The Minuteful Kidney Test shows high sensitivity, exceeding the guideline-required threshold (>85%, P<0.001), and robust specificity for detecting albuminuria in high-risk populations. This device offers potential to improve CKD screening and management and facilitate early intervention in primary care and community settings, where accurate rule-out capabilities are essential.

## Introduction

Urinary albumin to creatinine ratio (uACR) testing is central to the diagnosis, staging, and management of chronic kidney disease (CKD), yet testing rates remain short of guideline recommendations (1,2). Albuminuria is widely recognized as the key and early biomarker for CKD, with elevated levels indicating kidney damage and increased risk of adverse outcomes, including progression to end-stage renal disease (ESRD) and cardiovascular mortality (3,4,5).

Early detection of albuminuria is crucial for managing CKD effectively and preventing related complications. Traditional methods for measuring uACR often involve complex laboratory procedures, limiting accessibility and leading to low adherence to recommended guidelines. To address the global health challenges of CKD, particularly among patients with diabetes, hypertension, cardiovascular disease, and other comorbidities (6,7), innovative testing approaches are needed. The emergence of point-of-care testing (POCT) and home testing devices offers a promising solution, especially for ambulatory or low- to middle-income settings, where rapid and cost-effective diagnostics are crucial (8). The 2024 KDIGO guidelines (9), along with the ADA guidelines (10), recommend semi-quantitative POCT with ≥85% sensitivity when laboratory access is limited or when POCT offers advantages like convenience, immediate results, elimination of sample transportation, or reducing health disparities.

The Minuteful Kidney Test (MKT) is an FDA-cleared, home-based semi-quantitative device that detects urinary albumin, creatinine, and their ratio (uACR), providing a user-friendly option that enhances access to kidney health assessments. The MKT has the potential to improve early detection and guideline-concordant management of CKD through at-home testing. This comparative study evaluates the diagnostic accuracy of the MKT device against a standard laboratory quantitative method in at-risk populations for kidney disease.

## Materials and Methods

The study was conducted from January to March 2024, analyzing 615 urine samples arriving at an accredited laboratory in San Diego, California. Samples originated from outpatient settings like physician offices, nursing, and rehab facilities in economically diverse areas. Inclusion criteria encompassed urine samples from individuals with established medical conditions or risk factors for kidney damage, including diabetes, hypertension, cardiovascular disease, personal history of acute kidney injury, family history of CKD, inherited kidney diseases, early-stage CKD, dyslipidemia, obesity, autoimmune diseases, recurrent kidney infections, kidney malignancies and use of nephrotoxic medications. These were verified via Electronic Medical Records (EMR). Exclusion criteria were extreme urine pH (≤ 4.0 or ≥9.0), diluted samples (creatinine ≤ 30 mg/dL or SG ≤ 1.003), and samples containing preservatives. Fresh samples were processed within six days, frozen samples up to three months. Upon arrival, each sample was split into two vials and assigned a unique ID for blind parallel analysis using both methods. For semi-quantitative uACR we utilized the MKT Device, an in-vitro diagnostic, home-use urine analysis test system. For quantitative uACR we employed the Beckman Coulter AU 480 Analyzer, a clinical chemistry analyzer widely used in mid-volume hospitals and laboratories, with FDA-cleared reagents for albumin and creatinine: the ‘B38858 Olympus UALB (Urine/CSF Albumin) Reagent’, and the ‘OSR6178 Olympus Creatinine Reagent’. The Beckman Coulter AU 480 analyzer was chosen as the reference standard for this study due to its widespread use in clinical laboratories, FDA-cleared reagents for albumin and creatinine testing, and its proven accuracy and reliability in quantitative uACR measurement, aligning with CLIA standards for diagnostic validation. Both the index test (MKT) and the reference standard (Beckman Coulter AU 480) were performed in parallel, with all samples analyzed using both methods on the same day, to ensure consistency and minimize potential variability due to sample degradation or storage conditions. Performers of both tests were blinded to clinical information and the results of the other method until study completion. Tests were validated to CLIA standards. Albuminuria was categorized per KDIGO guidelines: A1 (Normal to mildly increased, uACR ≤ 30 mg/g) as negative, A2 (Moderately increased, uACR 30-300 mg/g) and A3 (Severely increased, uACR ≥ 300 mg/g) as positive. Samples were screened for integrity by pH, creatinine, urinalysis strips, and LC-MS (Agilent LC/MS 6410) for drug interferents. Concentrated samples were diluted as per standard laboratory practices. Data on sample age, storage conditions, visual inspection, collection methods and participant demographics—including gender, age, medical conditions, and chronic medications, were recorded to evaluate MKT’s performance across diverse clinical scenarios in at-risk populations. We followed STARD guidelines for diagnostic studies. Values and exclusion criteria were based on the Mandatory Guidelines for Federal Workplace Drug Testing Programs using Urine (11). Sensitivity, specificity, PPV, and NPV were calculated to evaluate the MKT device against the standard quantitative method. The projected NPV and PPV of the MKT were evaluated across a range of albuminuria prevalence rates using the observed sensitivity and specificity. Continuous variables were compared using t-tests; categorical variables with Chi-squared or Fisher’s exact tests. Statistical significance was set at p < 0.05 with 95% confidence intervals.

## Results

Of the 615 urine samples analyzed, 60 (9.8%) were excluded: 12 for extreme pH, 42 for dilution, and 6 for both conditions **(Figure 1)**. The final analysis included 555 samples, with 293 (52.8%) from female subjects. The mean age of study participants was 63.3 years (SD ±12.5), ranging from 24 to 101 years. Among the samples, 110 (19.8%) were from individuals with diabetes (type l or ll), 238 (42.9%) with hypertension, and 82 (14.8%) with cardiovascular disease. **(Table 1)**.

**Table 1.** Demographic and Clinical Characteristics of Study Population.

|  | **Overall**<br>n=555 (100%) | **Normoalbuminuria**<br>(A1) uACR ≤ 30<br>mg/g<br>n=417 (75.1%) | **Albuminuria**<br>(A2 -A3) uACR ≥ 30<br>mg/g<br>n=138 (24.9%) | **P-value** |
| --- | --- | --- | --- | --- |
| *Female vs.* | 293 (52.8%) | 221 (53.0%) | 72 (52.2%) | 0.944 |
| *Male* | 262 (47.2%) | 196 (47.0%) | 66 (47.8%) |  |
| *Age, years, mean (SD)* | 63.3 (12.5) | 62.3 (12.5) | 66.3 (12.1) | 0.001 |
| *Max age, years* | 101` | 101 | 98 |  |
| *Min age, years* | 24 | 29 | 24 |  |
| *Age ≥ 60* | 359 (64.7%) | 255 (61.2%) | 104 (75.4%) | 0.003 |
| *Diabetes\** | 110 (19.8%) | 66 (15.8%) | 44 (31.9%) | <0.0001 |
| *Hypertension* | 238 (42.9%) | 169 (40.5%) | 69 (50.0%) | 0.064 |
| *Cardiovascular Disease\*\** | 82 (14.8%) | 52 (12.5%) | 30 (21.7%) | 0.012 |
| *Kidney Disease †* | 23 (4.1%) | 14 (3.4%) | 9 (6.5%) | 0.171 |
| *Recurrent Urinary Tract*<br>*or Kidney Infections* | 34 (6.1%) | 22 (5.3%) | 12 (8.7%) | 0.212 |
| *Autoimmune Disease ‡* | 37 (6.7%) | 24 (5.8%) | 13 (9.4%) | 0.194 |
| *Nephrotoxic Medications*<br>*§* | 8 (1.4%) | 5 (1.2%) | 3 (2.2%) | 0.674 |
| *Dyslipidemia* | 206 (37.1%) | 149 (35.7%) | 57 (41.3%) | 0.283 |
| *Obesity* | 89 (16.0%) | 69 (16.5%) | 20 (14.5%) | 0.663 |
| *Tobacco use Disorder* | 132 (23.8%) | 93 (22.3%) | 39 (28.3%) | 0.190 |
| *Malignancies ¶* | 4 (0.7%) | 3 (0.7%) | 1 (0.7%) | 1.000 |

Data are presented as counts and percentages, or medians and ranges.
\* Type I or Type II Diabetes
\*\* Includes conditions such as Coronary Artery Disease (CAD), Heart Failure, and Peripheral Artery Disease (PAD)
† Includes conditions such as Acute Kidney Injury (AKI), Tubulointerstitial Nephritis, Medullary Cystic Kidney
Disease (MCKD), and known Chronic Kidney Disease (CKD).
‡ Includes conditions such as Systemic Lupus Erythematosus (SLE), Vasculitis, Gout, Rheumatoid Arthritis (RA),
Crohn's Disease, and Ulcerative Colitis (UC).
§ Includes medications such as Antivirals (e.g., Acyclovir, Valacyclovir), Methotrexate, and Lithium.
¶ Includes conditions such as Malignant Neoplasm of the Kidneys, Prostate, Bladder, Ureter, Multiple Myeloma, and
Leukemia.

**Figure 1.**
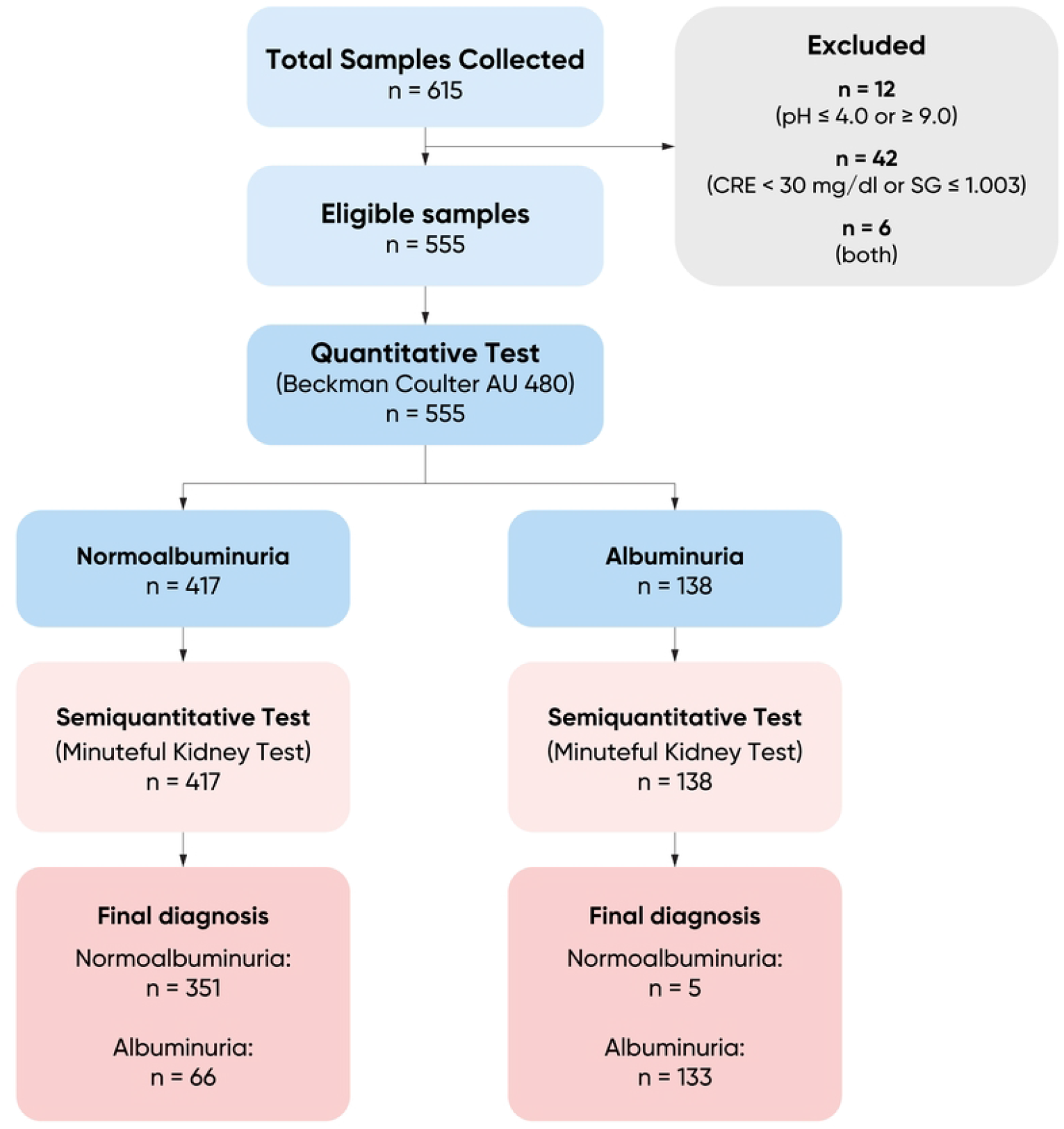
STARD Flow Diagram for Albuminuria Screening and Diagnosis

Albuminuria (uACR ≥30 mg/g) was identified in 24.9% of the samples using the quantitative standard device (95% CI 21.3-28.5). The semi-quantitative test showed a sensitivity of 96.4% (95% CI 93.3-99.5) and a specificity of 84.2% (95% CI 80.7-87.7). Regarding rule out and rule in utility, the NPV was 98.6% (95% CI 97.4-99.8) and PPV was 66.8% (95% CI 60.3-73.4). **(Table 2)**.

**Table 2.** Contingency Table: Minuteful Kidney Test vs. Beckman Coulter AU 480 analyzer for Albuminuria Detection.

|  | Beckman Coulter AU 480 quantitative analyzer |  |  | Total<br>(n) |  |
| --- | --- | --- | --- | --- | --- |
|  | Normoalbuminuria | Albuminuria |  |  |  |
| Minuteful Kidney<br>Semi-quantitative<br>Test | (A1) $\leq 30$ mg/g | (A2) 30-300 mg/g | (A3) $\geq 300$ mg/g | | |
| (A1) $\leq 30$ mg/g | 351 | 5 | | 356 | NPV<br>98.6% |
| (A2) 30-300 mg/g | 65 | 102 | 3 | 170 | PPV<br>66.8% |
| (A3) $\geq 300$ mg/g | 1 | 6 | 22 | 29 | |
| Total (n) | 417 | 113 | 25 | 555 |  |
| Positivity Rate 24.9% | Specificity 84.2% | Sensitivity 96.4% |  |  |  |

The MKT correctly classified 475 of 555 samples (85.6%) into the correct KDIGO albuminuria categories A1, A2, and A3, accurately identifying all 25 samples (100%) with a laboratory-confirmed A3 as abnormal. False negatives had uACR values from 32.4 to 57.5 mg/g, while false positives ranged from 3.9 to 28.7 mg/g **(Figure 2)**.

**Figure 2.**
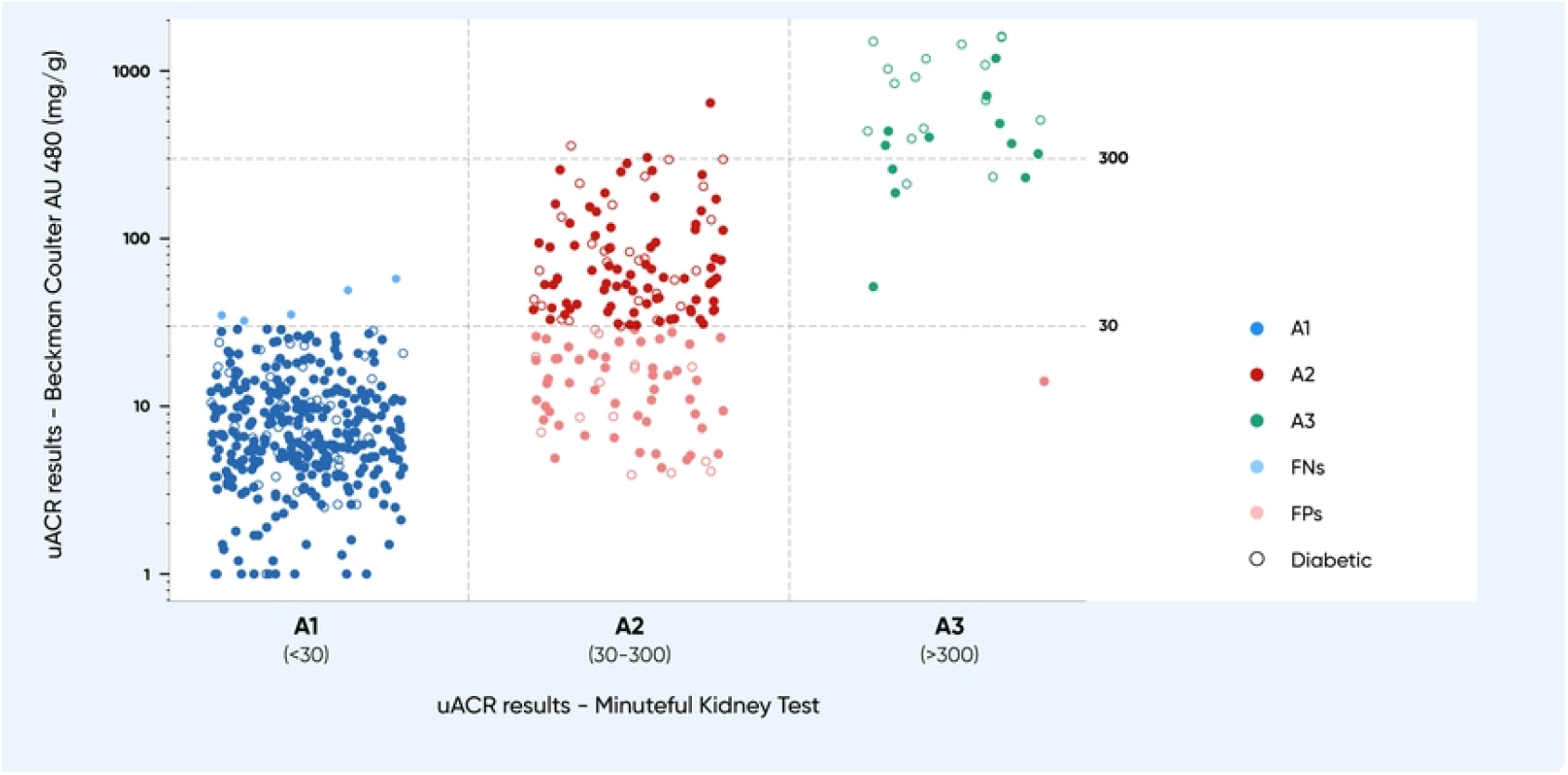
Scatter Plot: Minuteful Kidney Test vs. Beckman Coulter AU 480 Across KDIGO Albuminuria Categories

The analysis demonstrated consistent sensitivity across major CKD risk subgroups. Sensitivity was 100.0% (95% CI 90.0-100.0) in patients with diabetes compared to 94.7% (95% CI 87.5-98.0) in those without (P=0.18). For hypertension, sensitivity was 99.0% (95% CI 91.1-99.9) in patients with hypertension versus 94.2% (95% CI 85.1-98.1) in non-hypertensive individuals (P=0.37).

The projected NPV of the MKT consistently exceeds 90% even at very high albuminuria prevalence levels of up to 70%. The PPV of the test increases in settings with high albuminuria prevalence, such as populations at elevated risk for CKD **(Figure 3)**.

**Figure 3:**
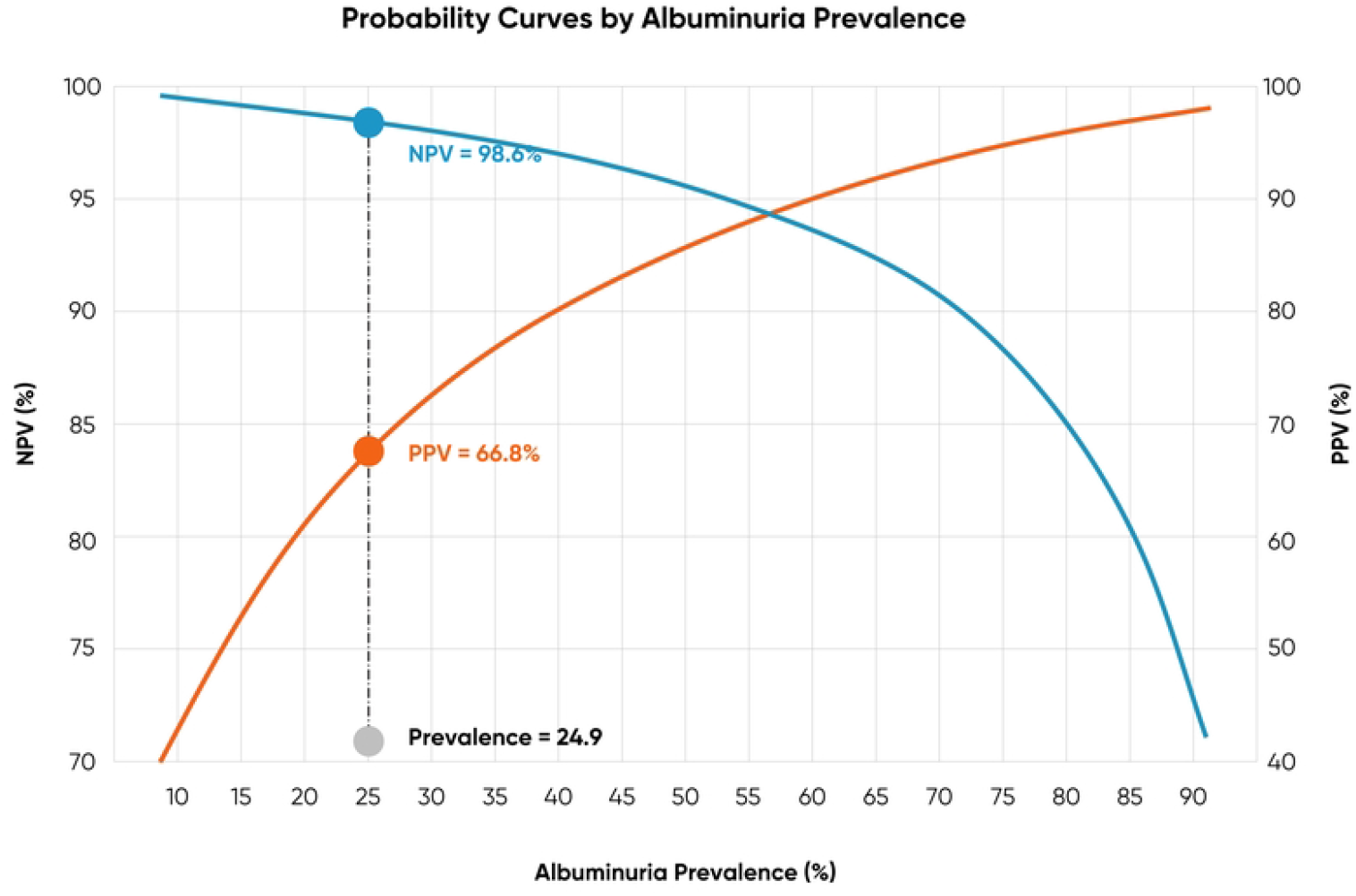
Probability Curves of Negative Predictive Value (NPV) and Positive Predictive Value (PPV) Across Different Albuminuria Prevalence Rates

## Discussion

Our findings show that the MKT’s high sensitivity (96.4%) and robust specificity exceed the 85% sensitivity threshold recommended by guidelines (P<0.001), reinforcing its clinical utility for early CKD detection and management in high-risk groups. The consistently high NPV across varying albuminuria prevalence levels ensures that individuals who test negative are highly likely to be free of albuminuria, supporting the MKT as a reliable screening tool in community and primary care settings, where ruling out CKD confidently is crucial. In high-prevalence settings, such as among high-risk populations, The MKT’s PPV increases from 40% to 98% as prevalence rises from 10% to 90%. This aligns with guidelines that recommend cautious interpretation and confirmatory testing of both quantitative and semi-quantitative positive results to account for the biological variability of albuminuria, reduce the risk of false positives, and promote accurate CKD diagnosis and management.

Although guidelines recommend CKD testing at least annually using both uACR and estimated glomerular filtration rate (eGFR), uACR testing remains suboptimal and inconsistent among at-risk populations, resulting in many undiagnosed cases. In addition, despite being the recommended standard, quantitative measurement of a first morning void sample faces challenges with standardization, including variability in testing methods and urine sample handling, which can affect results (13). Studies suggest that guideline-recommended uACR testing should increase CKD detection rates and improve patient care (12).

Our study highlights the clinical value of home and POCT methods like the MKT for assessing albuminuria. Expanding these approaches to population-wide screening has the potential to significantly improve testing rates, which have traditionally been low, and aligns with cost-effectiveness analyses and health policy recommendations advocating for enhanced CKD screening in at-risk populations (14). The MKT addresses this need by offering a user-friendly and accessible testing option that overcomes common barriers like sample transportation and the need for specialized laboratory equipment. It streamlines the diagnostic process in settings where rapid, low-cost testing is crucial, in line with KDIGO and ADA guidelines that recommend first morning void samples for their superior accuracy and reliability. The MKT facilitates convenient home testing of these samples, and future studies should quantify the reductions in false positives suggested by previous studies of first void urine testing (15,16).

Our study has some limitations. While semi-quantitative albuminuria categories provide actionable results comparable to quantitative tests, they cannot distinguish levels within KDIGO categories. The lack of uniform serum creatinine data prevented eGFR calculations and kidney failure risk assessments, limiting our ability to evaluate the predictive power of median ACR values across A1-A3 albuminuria, as seen with other dipsticks (17). We did not assess day-to-day biological variability or urine collection timing, though these factors are unlikely to affect accuracy metrics. Additionally, the study was confined to a single laboratory using the Beckman Coulter AU 480 Analyzer, which does not account for variations in the uACR reference method. Finally, while the MKT provides a critical step forward, follow-up testing and management by healthcare professionals remain essential to confirm diagnoses and implement appropriate treatment strategies effectively.

In conclusion, the MKT demonstrates high sensitivity and specificity, making it a valuable tool for implementing in U.S. clinical settings. Its use in home testing can help close gaps in current CKD screening and management guidelines.

## Data Availability

All data supporting the findings of this study are included within the manuscript and/or the Supporting Information files. Raw data from the study, encompassing both methods (the index device, Minuteful Kidney Test, and the comparator device, Beckman Coulter chemistry analyzer), are available upon request.

## Acknowledgments

Healthy.io funded the study.

